# Assessing the impact of reduced travel on exportation dynamics of novel coronavirus infection (COVID-19)

**DOI:** 10.1101/2020.02.14.20022897

**Authors:** Asami Anzai, Tetsuro Kobayashi, Natalie M Linton, Ryo Kinoshita, Katsuma Hayashi, Ayako Suzuki, Yichi Yang, Sung-mok Jung, Takeshi Miyama, Andrei R Akhmetzhanov, Hiroshi Nishiura

## Abstract

The impact of the drastic reduction in travel volume within mainland China in January and February 2020 was quantified with respect to reports of novel coronavirus (COVID-19) infections outside China. Data on confirmed cases diagnosed outside China were analyzed using statistical models to estimate the impact of travel reduction on three epidemiological outcome measures: (i) the number of exported cases, (ii) the probability of a major epidemic, and (iii) the time delay to a major epidemic. From 28 January to 7 February 2020, we estimated that 226 exported cases (95% confidence interval: 86, 449) were prevented, corresponding to a 70.4% reduction in incidence compared to the counterfactual scenario. The reduced probability of a major epidemic ranged from 7% to 20% in Japan, which resulted in a median time delay to a major epidemic of two days. Depending on the scenario, the estimated delay may be less than one day. As the delay is small, the decision to control travel volume through restrictions on freedom of movement should be balanced between the resulting estimated epidemiological impact and predicted economic fallout.

## 1. Introduction

By early February 2020, it was evident that the incidence of novel coronavirus infections (COVID-19) was exponential [1]. Accelerated by human migration, exported cases have been reported in various regions of the world, including Europe, Asia, North America, and Oceania [2]. To minimize the rapid growth of cases via human-to-human transmission [3-5], the government of China suspended all modes of transportation to and from Wuhan on 23 January 2020—including vehicles, trains, and flights—expecting that the intervention would prevent further spread of the disease [6]. As of 12 February 2020, two additional cities outside of Hubei Province—Wenzhou (Zhejiang Province) and Shenzhen (Guangdong Province)—have been placed on complete lockdown (i.e. no cross-border movement to and from the closed city) to prevent further spatial spread of COVID-19. To our knowledge, such drastic movement restrictions are a historical first.

Since Wuhan was placed on lockdown, travel restriction and border control have been implemented by various countries, either as: (i) complete travel bans, (ii) travel restriction and quarantine—which allows for restriction of healthy individuals, (iii) entry screening for all incoming travelers, or some combination thereof. Most countermeasures are in line with (ii) and (iii), aside from the three cities on complete lockdown, while some countries at high risk refused any entry from China (e.g. Australia) or those from Hubei province (e.g. Japan). All travel arrangements including tourist tours outbound from China (to international destinations) were cancelled, and all non-urgent travel with business purposes both inbound and outbound were greatly reduced.

The effectiveness of quarantine (i.e. lockdown) measures to prevent the spread of an epidemic due to a novel infectious pathogen where no vaccine is available has often been a subject of debate [7-9]. Under ordinary circumstances, border control efforts do not go beyond entry screening. However, during the epidemic of severe acute respiratory syndrome (SARS) in 2002–3, although entry screening at airports and other key locations was adopted in most countries its effectiveness was estimated to be very limited due to the relatively long incubation period and low prevalence of SARS, which resulted in extremely low positive predictive values at screening locations [10-13]. In the ongoing COVID-19 epidemic, many countries have accompanied regular entry screening with a drastic changes in travel restrictions. Although the effectiveness of entry screening is likely very limited as already shown elsewhere [14], the epidemiological impact of the change in movement restrictions has yet to be explicitly evaluated.

In this study, we quantify the impact of the drastic reduction in travel volume—resulting from movement restrictions—on the transmission dynamics outside China. We aim to estimate reductions in the number of exported cases, probability of an outbreak occurring outside China, and any time delay to a major epidemic that may be gained with these policies. We use the example of Japan, the country in Asia that receives the largest number of visitors from China, to calculate our estimates.

## 2. Methods

### 2.1. Epidemiological data

An epidemiological dataset of confirmed cases with COVID-19 infection diagnosed outside China was collected from government and news websites quoting official outbreak reports. For each case, the date of reporting and country of diagnosis were recorded. The data included only cases diagnosed outside China, but for whom infection may have occurred either in or outside China—our study focused on the latter. The dataset is available as a Supplementary Material (Table S1). All cases were confirmed using reverse transcriptase polymerase chain reaction (RT-PCR) apart from two cases in Australia that were clinically diagnosed. The endpoint for data collection was set at 6 February 2020.

### 2.2. Statistical model

We considered the impact of reduced travel volumes on COVID-19 transmission dynamics outside China. Specifically, we quantified the impact on: (i) the number of exported cases, (ii) the probability of a major epidemic, and (iii) the time delay to a major epidemic.

#### 2.2.1. Reduced number of exported cases

Figure 1 shows the observed number of infections in and outside China. The first exported case in Thailand was reported on 13 January 2020. Assuming the epidemic start date is set at 1 December 2019 (Day 0), the city of Wuhan was put in lockdown from Day 53 (or 23 January 2020). Considering that the mean incubation period of COVID-19 approximately 5 days, the impact of reduced travel volumes would start to be interpretable from Day 58 (28 January 2020). We used data from Day 43 (13 January) onwards. Because the first case diagnosed outside China was reported on that day.

**Figure 1.**
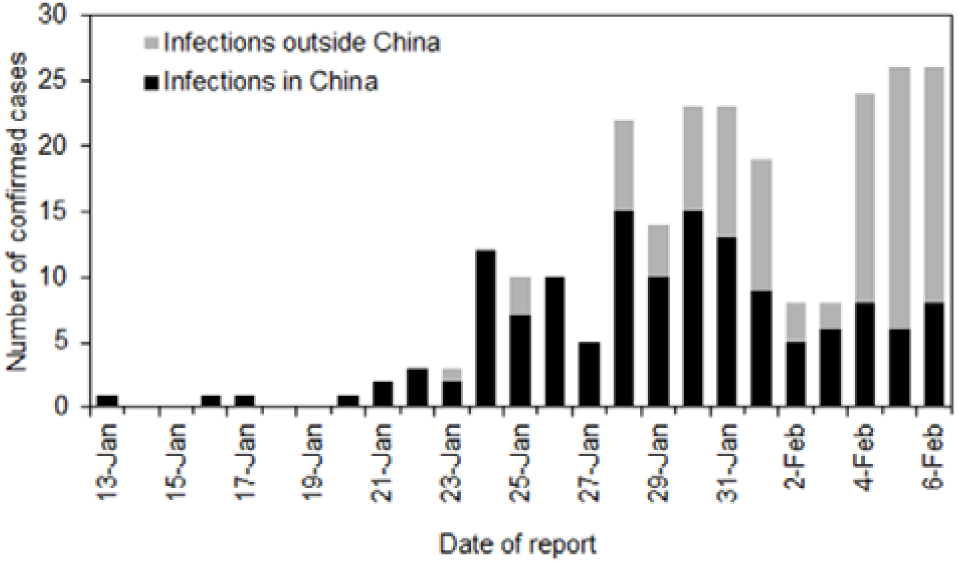
Number of confirmed cases outside China by date of report. The bars measure the number of cases reported each day between 13 January and 6 February 2020. The black bars represent infections that are likely to have occurred in China while the grey bars indicate infections that are likely to have occurred outside China.

To estimate the reduced volume of exported cases, we employ a counterfactual model. If we let c(t) be the incidence of exported cases on Day t, Poisson regression was used to fit the following model through Day 57:

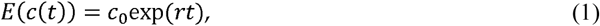

where *c*_0_ is the initial value at *t=*0 and *r* is the exponential growth rate of exported cases outside China. Using the estimated parameters and their covariance matrix, we obtain the expected number of exported cases from Day 58 onwards. Supposing that *h*(*t*) is the observed number of cases on day *t*, the reduced travel volume of exported cases by Day 67 is calculated as:

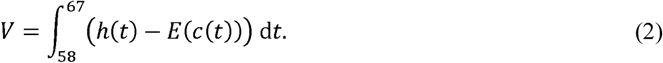

#### 2.2.2. Reduced probability of a major epidemic overseas

We assumed that the distribution of the number of secondary cases generated by a single primary case follows a negative binomial distribution with the basic reproduction number *R*_0_, i.e., the average number of secondary cases generated by a single primary case, and the dispersion parameter *k*. The probability of extinction *π* is then modeled as:

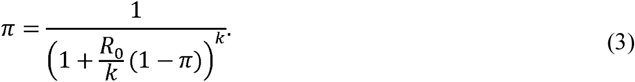

*R*_0_ is estimated to range from 1.5 to 3.7, and here we adopt 1.5, 2.2 and 3.7 as plausible values for our calculations [15-17]. The value of *k*, a dispersion parameter, is assumed to be 0.54 as estimated elsewhere [16].

Supposing that there are *n* untraced cases that were independently introduced, the probability of a major epidemic is:

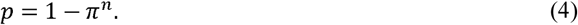

Now we compare two scenarios: the observed data as influenced by the reduction in travel volume, and a counterfactual scenario in which travel volume reduction does not take place. *m* describes the cumulative number of exported COVID-19 cases observed in the former scenario, while 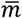 describes the number of cases in the counterfactual scenario. This leads to the following integrals:

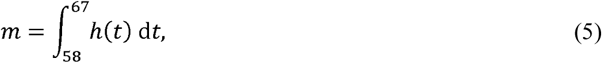

and

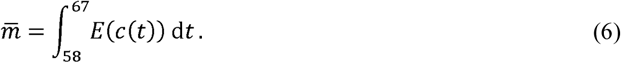

Accordingly, the reduced probability of a major epidemic is calculated as:

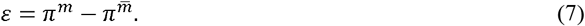

It should be noted that the probability of a major epidemic is evaluated at the country level, and only results for Japan are presented here as equally handling the probability of a major epidemic for each importing country is too challenging. For the computation, we first subtracted 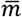, the integral of *E*(*c*(*t*)), by the integral of *h*(*t*), assuming that all cases *h*(*t*) were already traced, and then we multiplied the difference by 0.9, 0.7, or 0.5 if only 10%, 30%, or 50% of contacts were traced, respectively. For *m*, we accounted for three symptomatic cases that were regarded as locally acquired infections in reports and diagnosed between Day 58 and Day 67. Assuming that the asymptomatic ratio was 50% [18], we assumed that in total there were *m*=6 untraced cases including the diagnosed cases.

#### 2.2.3. Time delay to a major epidemic gained from the reduction in travel volume

Lastly, we measured the time delay to a major epidemic gained from the reduction in travel volume using the hazard function of a major epidemic, *λ*(*t*), in the absence of travel volume changes. We model the probability of a major epidemic by time *t* in the absence of travel volume reduction as follows:

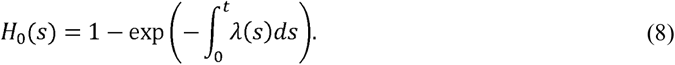

In the presence of travel volume reduction, the hazard is reduced by the relative reduction factor in the probability of a major epidemic:

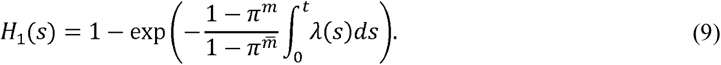

Consider the median time to a major epidemic in (8) and (9). Since an exponential growth of cases has been observed, we let the hazard be an exponential function. Then, the integral of the hazard function holds the form: *C* · (*exp*(*rt*) − 1) where *C* is a constant (assumed to be one for the following calculation), and *r* is the exponential growth rate estimated at 0.14 per day [15]. The doubling time is then calculated as *t*_d_=ln(2)/*r*=4.95 days. The difference in the median date between (8) and (9) is thus described as:

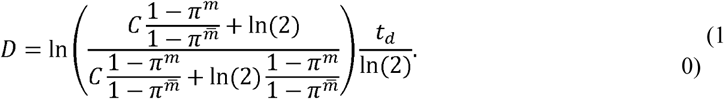

All computations were conducted in JMP version 14.0 (SAS Institute, Cary, North Carolina).

## 3. Results

In Figure 1, a total of 242 cases were diagnosed and reported outside of China in 27 countries between Day 43 (13 January) and Day 67 (6 February 2020). Of these, 140 cases were considered to have been infected in China and 102 cases were considered to have been infected outside China. The country with the highest number of exported cases infected inside China was Thailand (n=20), followed by Singapore (n=18), Australia (n=14), and Japan (n=12). Among 242 cases, we specifically focus on 140 cases who traveled while movement restrictions were in place and were likely affected by said restrictions. Figure 2 compares the observed and expected number of cases diagnosed outside China by date of report. The exponential growth of cases is consistent with the exponential growth of incidence in China, which qualitatively captures the observed pattern of incidence from Day 43 to Day 57. Using the predicted curve, the expected number of cases between Day 58 (28 January 2020) and Day 67 was 321 cases (95% confidence interval [CI]: 181, 544). In the empirical observation, a total of 95 cases were diagnosed, including 8 cases in Japan. That is, following the time that Wuhan city was put in lockdown, we estimate that 226 cases (95% CI: 86, 449) were prevented from being exported across the world. This corresponds to a reduction in the number of exported cases of 70.4% during that time period.

**Figure 2.**
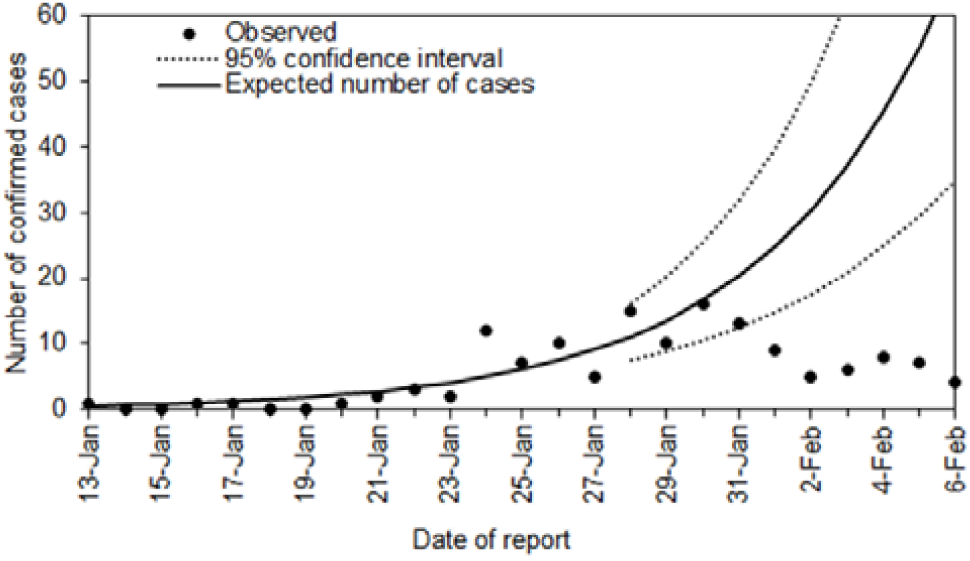
Observed and expected number of cases diagnosed outside China by date of report. Observed cases (dots) include those infected in China. An exponential growth curve was fitted to the observed data from 27 January 2020. The dashed lines represent the 95% confidence interval on and after 28 January 2020.

As another measure of impact, we estimated the probability of a major epidemic. Figure 3A shows the probability of a major epidemic with three different levels of transmissibility assuming an R0 of 1.5, 2.2, or 3.7, and three different levels of contact tracing resulting in a success rate of isolation of the traced contacts of 10%, 30%, or 50%. Without the reduction in the travel volume, the probability of a major epidemic exceeded 90% in most scenarios. However, considering there have been six untraced cases in Japan under travel restrictions, the probability of a major epidemic more broadly ranged from 56% to 98%. Figure 3B shows the reduced probability of a major epidemic. Assuming an R0 of 2.2, the absolute risk reduction was 7%, 12%, and 20%, respectively, for contact tracing levels leading to isolation at 10%, 30%, and 50%.

**Figure 3.**
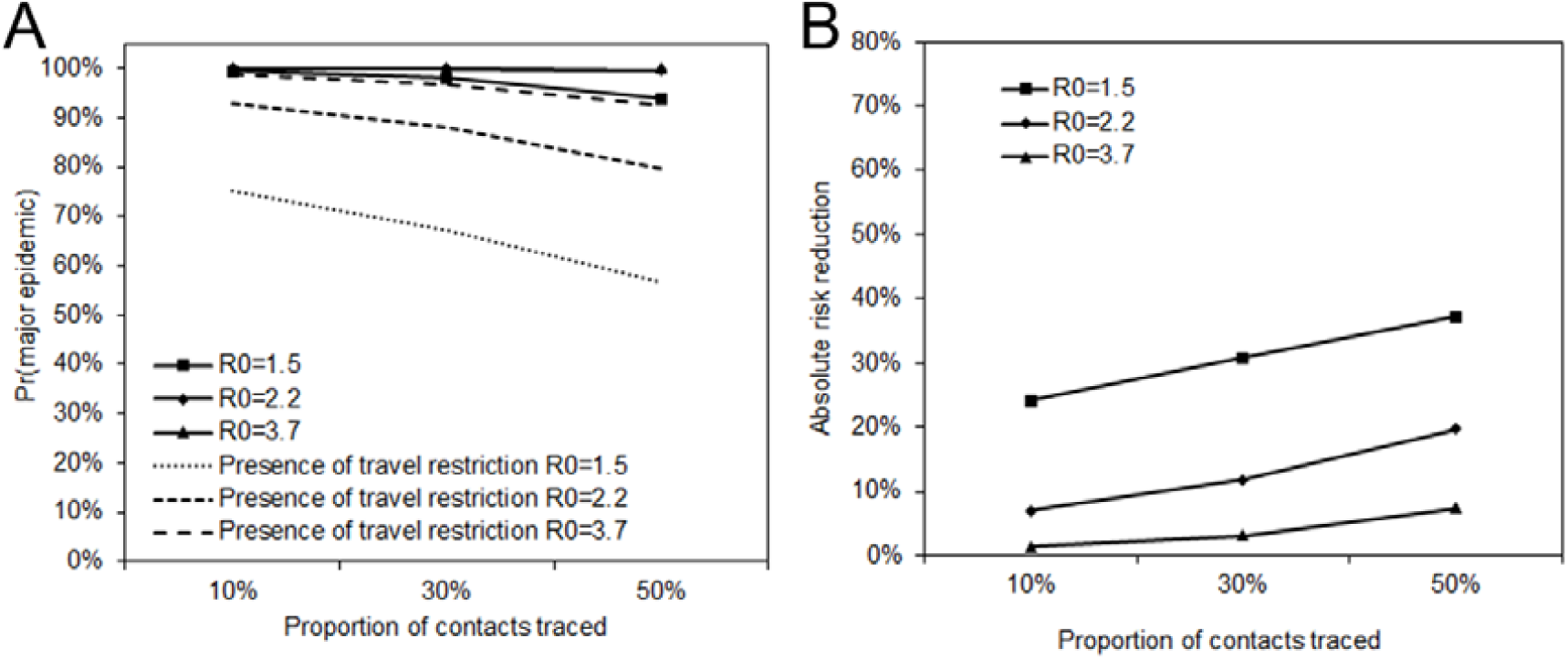
Probability of a major epidemic with various levels of transmissibility and traced contact. (A) The solid lines represent the probability of a major epidemic in the counterfactual scenario, i.e., based on the expected number of cases diagnosed in Japan. Dashed lines represent the probability of a major epidemic in the presence of travel volume reductions, calculated using the number of traced and untraced cases was 6 in total in Japan from Day 58 to Day 67. Contact tracing leading to isolation was assumed at three different levels: 10%, 30%, and 50%. (B) The vertical axis represents the reduced probability of a major epidemic due to travel volume reduction. The horizontal axis shows the proportion of cases traced, adopting the same scenarios as panel A.

Figure 3B describes the absolute reduction in risk of a major epidemic. The largest reduction was 37% when *R*_0_ =1.5 and 50% of contacts were traced. The smallest reduction was 1% when *R*_0_ =3.7 and 10% of contacts were traced. Using those estimated relative reductions, the median time of delay gained by travel volume reduction is shown in Figure 4. The time delay of a major epidemic was less than one day when *R*_0_ is 2.2 and 3.7, and 1 to 2 days when *R*_0_ is 1.5.

**Figure 4.**
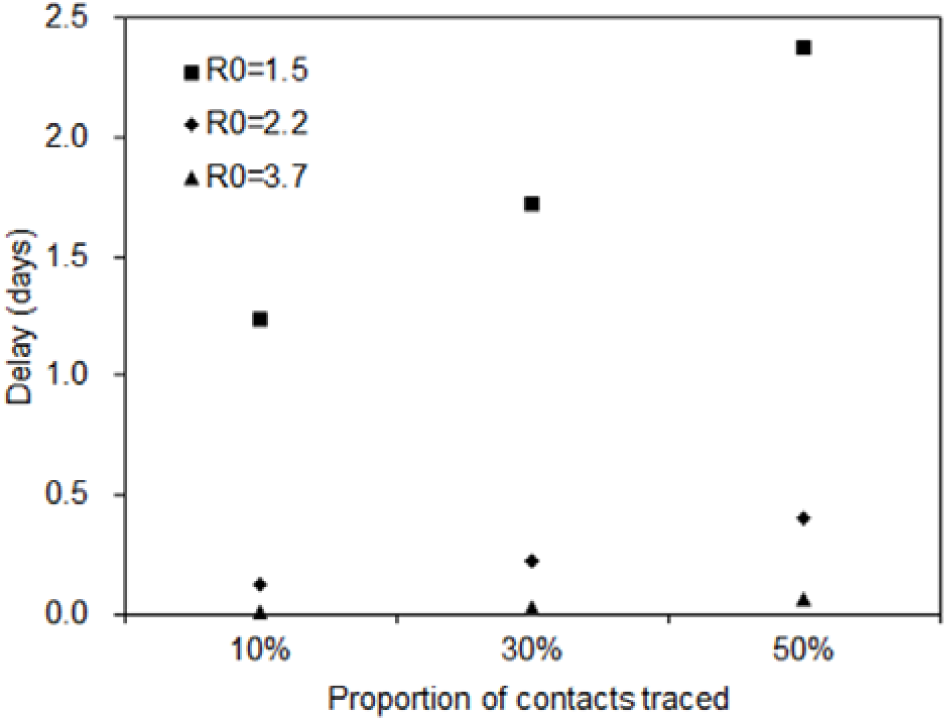
Delay in the time to a major epidemic gained by travel volume reduction. The median delay is shown for Japan, using relative reduction in the probability of a major epidemic. The vertical axis represents the time delay to a major epidemic (in days), and the horizontal axis represents the proportion of contacts traced. Each shaped dot represents different values of the basic reproduction number.

## 4. Discussion

The present study explicitly quantified the epidemiological impact of reduced travel volume to and from China on the transmission dynamics of COVID-19 outside China using simple statistical models. The three epidemiological outcomes we measured were: (i) the number of exported cases, (ii) the probability of a major epidemic, and (iii) the time delay to a major epidemic. When the volume of exported cases outside China was considered to have been reduced by 70.4% the probability of a major epidemic was estimated to be reduced by 7–20% in Japan, and 2-day delay was gained in the estimated time to a major epidemic between Day 58 and Day 67.

The reduced volume of exported cases was estimated to be as large as 226 cases outside China. Our estimate is consistent with an assessment by Chinazzi et al. [19], which indicated that the exported cases would be reduced by 80% by the end of February. In addition to appropriately quantifying the impact on prevention of exported cases, we have estimated the median time delay to a major epidemic assuming plausible values of R0 at 1.5, 2.2, and 3.7. With reduced probability of a major epidemic, the time delay to a major epidemic was estimated at a maximum of 2 days in Japan and a minimum of less than 1 day. The estimated effect of the delay to a major epidemic outside China is smaller than what was anticipated for cities in China other than Wuhan. Tian et al. [20] estimated that the reduction in travel volume led to a 2.9-day delay in the spatial spread in China. To our knowledge, the present study is the first to have used simple stochastic process models to explicitly estimate the time delay to a major epidemic in Japan that gained by the drastic reduction in travel volume in and outside China.

Although the COVID-19 epidemic was declared a public health emergency of international concern (PHEIC) by the World Health Organization (WHO), the WHO specifically called upon member states to not restrict the freedom of movement of persons as a result of the epidemic [21]. However, member states did not adhere to this recommendation and have varyingly restricted the free movement of people from China [22]. Such restrictions were most drastic in China, where some cities were put on complete lockdown [22]. These political decisions regarding movement restrictions must balance the expected epidemiological impact with predicted economic burden—the latter of which we did not examine. While securing a few days delay to epidemic spread in China would secure time for healthcare systems in Chinese cities that have not yet been affected to prepare for the appearance of case-patients [20], the impact of such a delay outside China is not substantial enough to accomplish meaningful prevention, such as the development, manufacturing, and distribution of a vaccine.

In modern history, this epidemic is perhaps the first instance where a large city populated with more than 10 million people was placed on lockdown. While countermeasures to prevent epidemic spread require the sort of strong political decisions that resulted in strong movement restrictions, our study indicated that the delay to a major epidemic in countries other than China (using Japan as an example) was unfortunately minimal. While the complete lockdown of Wuhan, Wenzhou, and Shenzhen likely helped reduce case incidence outside of these cities, migration from other cities in China could still contribute to the spread of infection locally and internationally [23]. To quantify the epidemiological impact for the entire course of the epidemic more precisely, a more detailed analysis using dynamic datasets, e.g. airline passenger data, should be explored in the future.

Limitations of the present study must be discussed. First, the present study relied on the volume of cases diagnosed outside China and did not directly examine human migration data. Second, we were unable to classify exported cases into those who acquired infection in Hubei versus elsewhere in China. Having this information may offer additional insight. Third, several rough assumptions (e.g., a fixed time delay from illness onset to reporting at 5 days) were imposed, and the results presented here should be regarded as a preliminary assessment.

In conclusion, the present study explored the impact of reduced travel volume to and from China on the transmission dynamics of COVID-19 outside China, estimating that the time delay to a major epidemic was on the order of 2 days by 7 February 2020 for Japan. Our proposed approach was kept simple and will be applicable to other emerging epidemics in the future.

## Data Availability

The dataset is available as a Supplementary Material (Table S1).

## Supplementary material

The following is available online at www.mdpi.com/xxx/s1, Table S1: Number of confirmed cases by date of report.

## Author Contributions

H.N. conceived the study, A.A. and T.K. collected the data and analyzed the empirical data using models. All authors participated in the study design. A.A, T.K. and H.N. drafted the manuscript. All authors gave comments on the earlier versions of the manuscript. All authors edited the manuscript and approved the final version.

